# A Positive Response around 4.6 ms in Auditory Brainstem Response in Cochlear Nerve Deficiency Infants

**DOI:** 10.1101/2023.01.05.22283540

**Authors:** Bei Li, Meiping Huang, Mengda Jiang, Wentao Shi, Hao Wu, Zhiwu Huang, Yun Li

## Abstract

**Objectives:** A positive response around 4.6 ms latency (IV’ wave) was observed at high intensity level in the ears which were always diagnosed with cochlear nerve deficiency (CND). This study is aimed to investigate the relationship between the IV’ wave and CND.

**Study Design:** Retrospective study.

**Setting:** Tertiary hospital

**Methods:** The raw Auditory Brainstem Response data and inner ear images of the infants were reviewed. Data were analyzed by ear and the CND ears were further divided into the IV’ wave present and IV’ wave absent group. The distortion product otoacoustic emissions (DPOAE) and auditory steady-state response (ASSR) results were compared between the two groups.

**Results:** In total, 570 ears were included. There were 129 ears diagnosed with CND and the IV’ wave was observed in 52 ears. The latency of the IV’ wave averaged 4.60 ms at 95 dB. The positive predictive value of the IV’ wave for CND was 98.1%. The incidence of the IV’ wave in the CND is highly unlikely to occur by chance. The differences of the DPOAE amplitude between the two groups were not significant. The ASSR results of the IV’ wave present group was significantly better.

**Conclusions:** Our study demonstrated a 4.6 ms positive response at high intensity level in the infants. The IV’ wave showed an excellent positive predictive value for CND. The extrapolated thresholds of the IV’ wave present group were better. The IV’ wave is expected to be a new indicator for the CND infants.

## INTRODUCTION

Auditory neuropathy spectrum disorder (ANSD) is thought to account for 7–10% of all childhood permanent hearing impairment^1^. It is an audiological diagnosis typified by the presence of intact outer hair cell function in the cochlea, reflected by the presence of otoacoustic emissions or cochlear microphonics, together with severely abnormal or absent auditory brainstem responses. The pathogenesis of ANSD encompasses a wide range of disease mechanisms, with pathologies localized to multiple sites along the auditory pathway, including inner hair cells^2^, synapses^3^, spiral ganglion neurons^4^, auditory nerve^5^, or brainstem auditory nuclei^6^. Different etiologies are associated with different audiologic features, and auditory steady-state response (ASSR) was suggested to serve as an objective measure for estimating behavioral thresholds in ANSD patients^7^.

Cochlear nerve deficiency (CND) represents a ‘‘literal’’ or true form of ANSD. It is diagnosed by inner ear imaging examination, including high-resolution computed tomography (HRCT) and magnetic resonance imaging (MRI). Although HRCT cannot visualize nerves, it can resolve the bony cochlear nerve canal (BCNC), which has also been reported to be narrower on average in ears with CN aplasia^8–10^. Because of the cochlear nerve aplasia or hypoplasia, the CND population would have disordered neural transduction and might share some audiological features with other ANSD.

During the last two decades more patients with CND have undergone cochlear implantation (CI). Researches showed that children with CND can still benefit from CI^11–13^. However, the reported audiologic outcome in children diagnosed with CND is poorer and more variable than those without nerve deficiency^12,14^. It was suggested that expected benefit can be dependent on the status of the CN. As a result, researchers have looking for the predictive role of the status of the cochlear nerve in radiographic images for CI outcomes in CND patients, such as the diameter of the internal auditory canal^12,15^, internal auditory meatus nerve grading system^11^, number of nerve bundles and vestibulocochlear nerve area^16^. Kari et al. concluded that current imaging modalities cannot accurately depict the status of the cochleovestibular nerve or predict a child’s benefit with a CI^15^.

In our center, a positive response at approximately 4.6 ms latency (IV’ wave) was observed at high intensity level (80 to 95 dB) while the regular V wave was absent in the auditory brainstem response (ABR) tests in some referred infants. In the further inner ear imaging examinations, those infants were always diagnosed with CND in the ear that presented the IV’ wave. In order to study the relationship between the IV’ wave and CND, a retrospective study was designed. The air-conducted click ABR waveforms were reviewed and the image information of all the individuals presenting the IV’ wave was listed. A chi-squared test was applied. The profile of the IV’ wave was summarized, including latency and intensity-incidence. The CND ears were further divided into the IV’ wave present and IV’ wave absent group and the distortion product otoacoustic emissions (DPOAE) and ASSR results were further compared between the two groups.

## METHODS

This was a retrospective study, and it was approved by the Translational Medicine Ethics Review Committee of Shanghai Ninth People’s Hospital Affiliated to Shanghai Jiao Tong University, School of Medicine (SH9H-2022-T378-1). The raw data of air-conducted click ABR waveforms of all infants who visited our center between February 2017 and December 2021 were reviewed. The inclusion criteria included profound hearing loss (air-conducted ABR threshold > 95 dB) on at least one side. The exclusion criteria included age older than 36 months, tympanogram type B, specific ear trauma and infection history or other acquired hearing loss disease, and no imaging examinations of the inner ear taken in our hospital.

Firstly, 877 ABR recordings that met the criteria were screened out, of which 46 recordings were found follow-up recordings. In total, there were 831 patients. Thirty-five patients were excluded for a first visit age older than 36 months; twenty-nine patients were excluded for tympanogram type B and 412 patients were excluded for lack of imaging examination in our hospital. Finally, 355 patients were included. For those patients who had follow-up visits, only the recordings of the first visit were counted.

In the 355 patients, 140 individuals had profound hearing loss in one ear, of which 80 showed profound hearing loss in the left ear and 60 showed profound hearing loss in the right ear. 215 individuals had profound hearing loss in both ears. In total, 570 ears were included. 140 individuals only underwent HRCT, 46 individuals only underwent MRI and 169 individuals underwent both examinations.

Imaging results were based on reports reviewed by radiologists with more than five years of experience. The diagnostic criteria for CND are CN nerve smaller than the facial nerve on oblique sagittal images in the MRI or BCNC ≤1.5 mm in the HRCT.

All the audiological assessments of the individuals including air-conducted click ABR threshold, DPOAE, and ASSR were reviewed. The DPOAE amplitude were calculated by averaging the absolute amplitude in dB SPL of DPOAEs in the ear across all frequencies (1–9 kHz). The extrapolated threshold of frequency 500, 1000, 2000 and 4000 Hz was measured by the ASSR. If there was no response at the maximum output of the audiometer, the extrapolated threshold was considered 5 dB greater than the maximum output for the purpose of statistics. The details of the protocol and parameters of the ABR, DPOAE and ASSR tests were displayed in Supplementary I.

The features of the IV’ wave was summarized, including latency and intensity-incidence (threshold). The relationship of the IV’ wave and CND was calculated by a chi-squared test. All the data were analyzed by ear and were further grouped by IV’ wave and CND diagnosis. The positive predictive value of the IV’ wave for CND was calculated. The DPOAEs and ASSR results were also compared between the IV’ wave present and absent groups.

## Results

### The profile of the IV’ wave

The age of the individuals observed with the IV’ wave ranged from 3 to 27 months. The IV’ wave amplitude dropped significantly as the sound intensity decreased to 85 dB. The latency of the IV’ wave was barely prolonged as the sound intensity decreased. In the 570 ears, the IV’ wave was observed in 52 ears of 40 individuals. Twenty-four ears were observed on both sides of 12 individuals with profound bilateral hearing loss. Seven ears were observed on only one side of 7 individuals with profound bilateral hearing loss. Twenty-one ears were observed in 21 individuals with unilateral profound hearing loss. The IV’ wave of 3 cases are shown in Figure 1.

**FIGURE 1.**
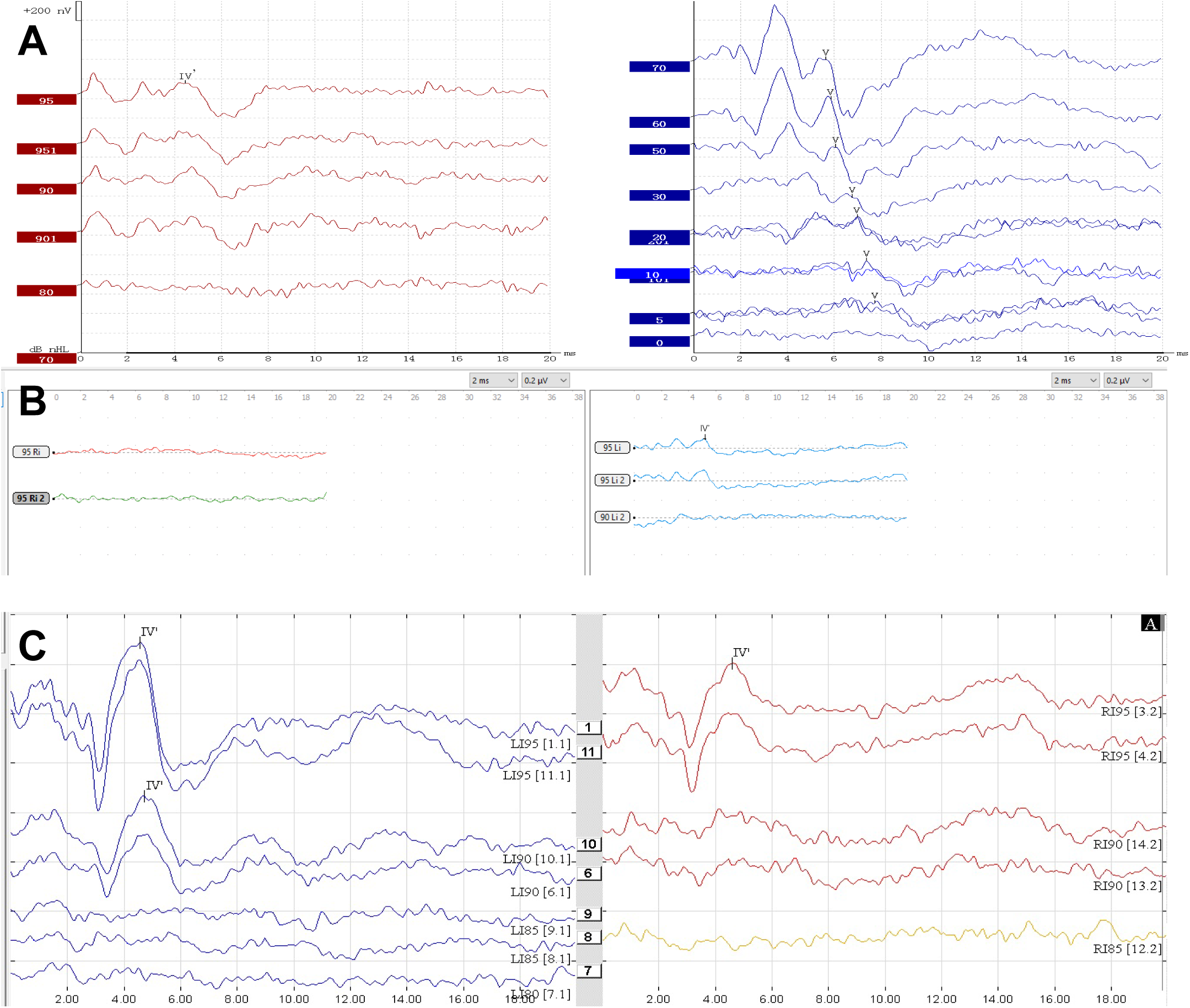
Examples of the IV’ wave in 3 cases. A, Normal V wave presents in the left ear and the IV’ wave presents in the right ear. B, The IV’ wave presents in the left ear and no wave presents in the right ear. C, The IV’ wave presents bilaterally

The latency and the intensity-incidence of the IV’ wave was calculated among the first visit ABR recordings in the 52 ears. The latency of the IV’ wave ranged from 3.87 to 5.19 ms, averaging 4.60 ± 0.314 ms at 95 dB. The intensity-incidence of the IV’ wave was calculated for threshold estimation. At 95 dB, 100% of ears displayed the IV’ wave (52/52). At 90 dB, 63% of ears displayed the IV’ wave (33/52). At 85 dB, 32% of ears displayed the IV’ wave (6/19). At 80 dB, 10% of ears displayed the IV’ wave (3/29). At 70 dB, none of the ears displayed the IV’ wave (0/2).

### The relationship between the IV’ wave and CND

Of the 52 ears that presented with the IV’ wave, 51 ears were all diagnosed with CND. The remaining one ear was diagnosed with large vestibular aqueduct syndrome accompanied by Mondini deformity. There are 78 ears diagnosed with CND and showed no IV’ wave. The positive predictive value of the IV’ wave for CND was 98.1%. The sensitivity of the IV’ wave for identifying CND was 39.5%. In the no CND group, there were 347 ears showed no inner ear malformation, 40 ears showed enlarged vestibular aqueduct and 53 ears showed cochlear or vestibular malformation including 16 ears Michel deformity. A chi-squared test employing a 2 x 2 contingency table showed that the incidence of the IV’ wave in the CND is highly unlikely to occur by chance (p < 0.001).

The information and imaging examination results of the individuals presenting with the IV’ wave is listed in Table 1. The CT and MRI images of the normal and abnormal inner ears are displayed in Figures 2 and 3, respectively.

**FIGURE 2.**
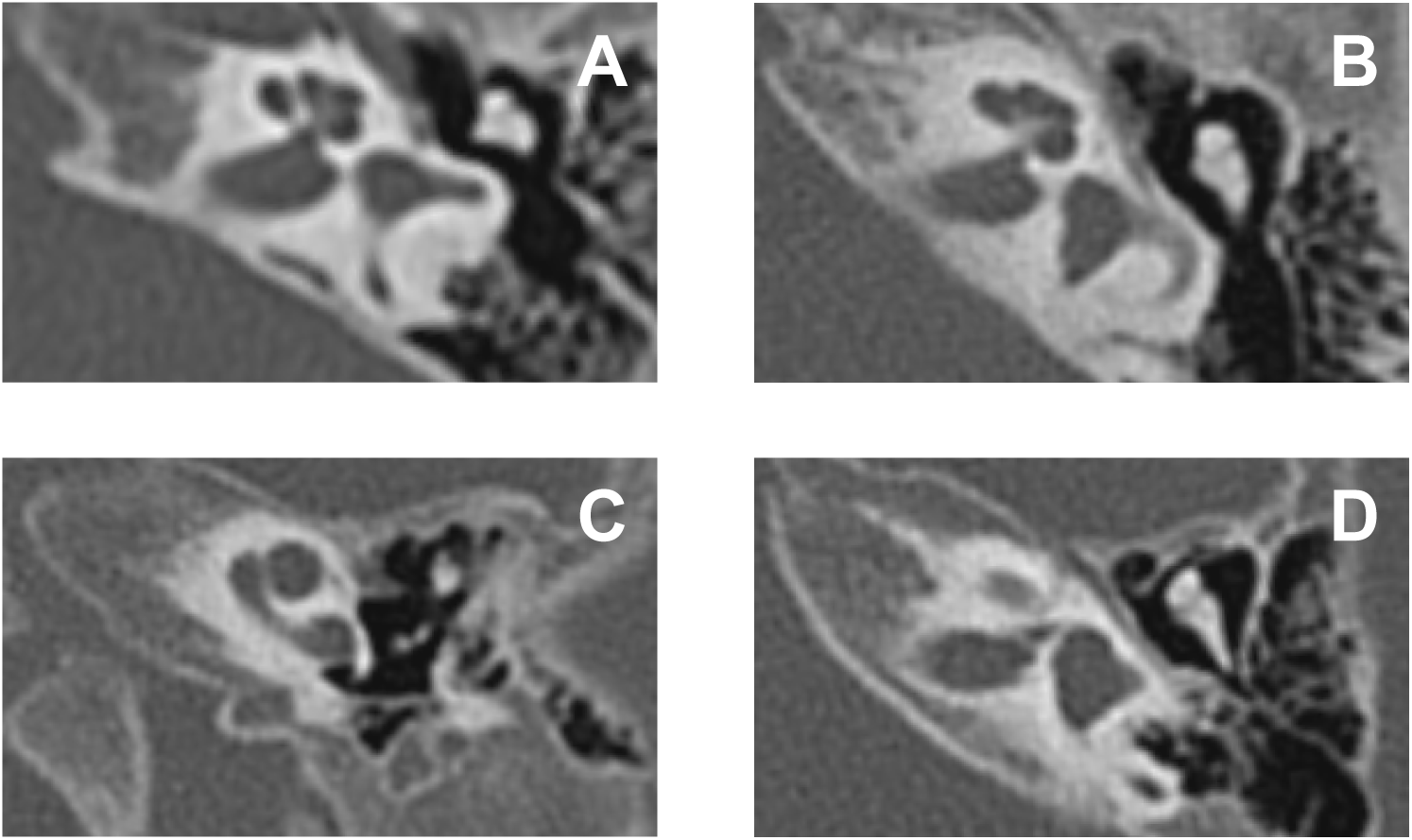
The normal and abnormal inner ear on high-resolution computed tomography. A, Normal inner ear imaging with profound hearing loss of the left ear (IV’ wave absent). B, Bony cochlear nerve canal stenosis with profound hearing loss of the left ear (IV’ wave present). C, Incomplete partition type I and enlarged vestibular aqueduct with profound hearing loss of the left ear (IV’ wave present). D, Bony cochlear nerve canal stenosis and vestibular semicircular canal deformity with profound hearing loss of the left ear (IV’ wave present)

**FIGURE 3.**
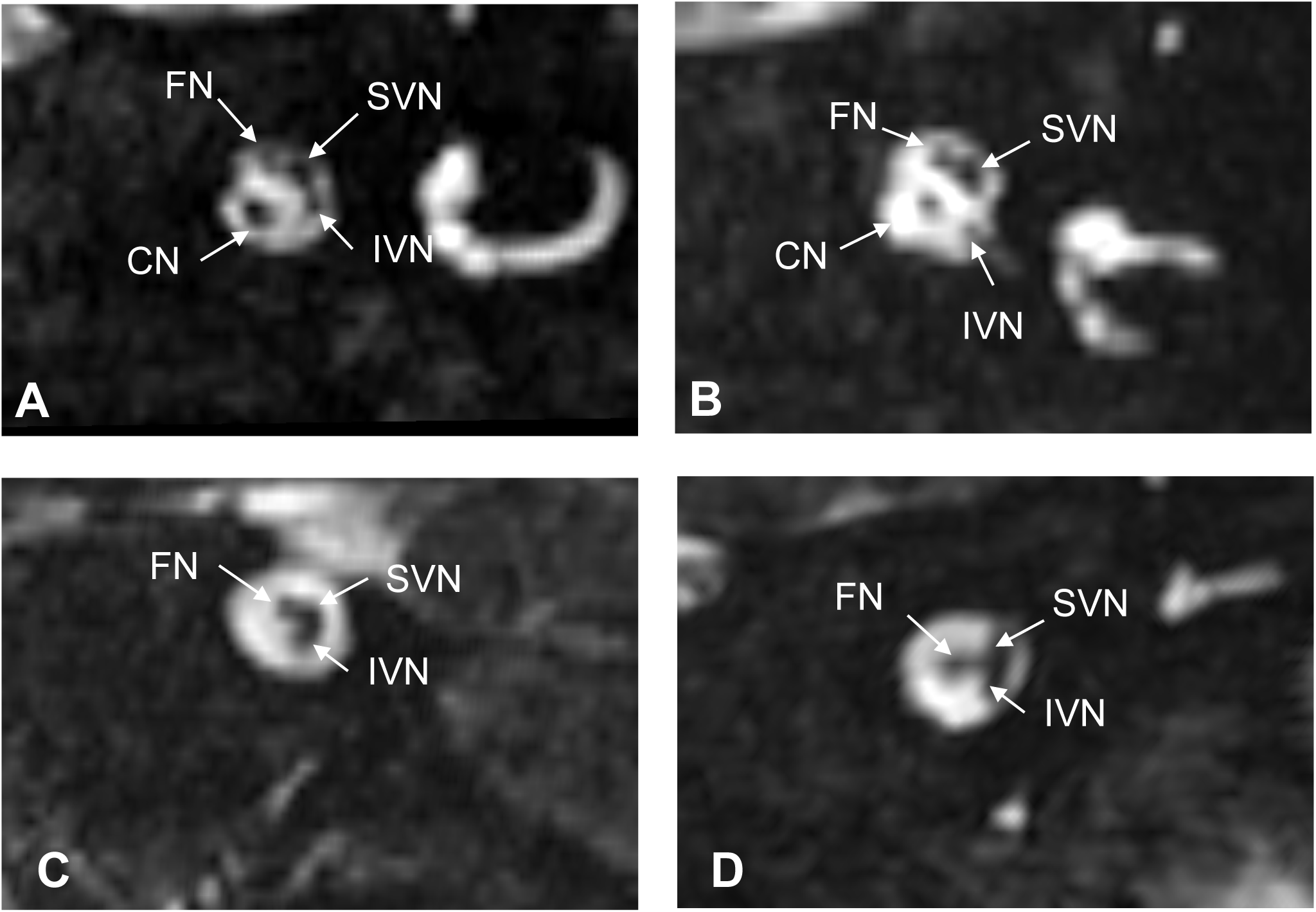
The normal and abnormal cochlear nerve on oblique sagittal magnetic resonance imaging. A, Normal cochlear nerve image with profound left ear hearing loss (IV’ wave absent). B, Cochlear nerve hypoplasia presenting IV’ wave with profound left ear hearing loss. C, Cochlear nerve absent without IV’ wave with profound left ear hearing loss. D, Cochlear nerve absent presenting IV’ wave with profound left ear hearing loss. CN: cochlear nerve; FN: facial nerve; IVN: inferior vestibular nerve; SVN: superior vestibular nerve

**Table 1.** The information and imaging examination results of the individuals presenting with the IV’ wave.

| Age(months)/sex | PHL side<br>/ IV' wave side | Inner ear malformations |  | CN on MRI |  |
| --- | --- | --- | --- | --- | --- |
|  |  | R | L | R | L |
| 3-6/M | B/B | - | - | Hypoplasia | Hypoplasia |
| 3-6/M | L/L | / | BCNCS | - | - |
| 3-6/M | L/L | / | BCNCS | - | - |
| 3-6/M | L/L | / | BCNCS | - | - |
| 3-6/M | L/L | / | BCNCS | - | - |
| 3-6/F | B/B | - | - | Absent | Absent |
| 3-6/F | L/L | / | BCNCS | / | Absent |
| 3-6/M | R/R | BCNCS | / | - | - |
| 3-6/F | L/L | / | BCNCS | / | Absent |
| 3-6/M | B/L | / | BCNCS | / | Absent |
| 3-6/F | B/B | BCNCS | BCNCS | Absent | Absent |
| 3-6/M | L/L | / | IP-I, enlarged vestibular<br>aqueduct | / | Hypoplasia |
| 3-6/F | B/B | BCNCS | BCNCS | Absent | Absent |
| 3-6/F | B/B | BCNCS | BCNCS | Absent | Absent |
| 3-6/M | B/B | BCNCS | BCNCS | Hypoplasia | Hypoplasia |
| 3-6/M | L/L | / | BCNCS | - | - |
| 3-6/M | B/B | BCNCS | BCNCS | Absent | Absent |
| 7-12/M | B/R | BCNCS | BCNCS | - | - |
| 7-12/M | B/L | BCNCS | BCNCS | - | - |
| 7-12/F | B/B | BCNCS | BCNCS | Absent | Absent |
| 7-12/F | R/R | BCNCS | / | - | - |
| 7-12/F | L/L | - | - | / | Hypoplasia |
| 7-12/M | B/L | BCNCS and Vestibular<br>semicircular canal<br>deformity | BCNCS and Vestibular<br>semicircular canal<br>deformity | - | - |
| 7-12/F | L/L | / | BCNCS | - | - |
| 7-12/F | L/L | / | BCNCS | / | Absent |
| 7-12/F | R/R | - | - | Hypoplasia | / |
| 7-12/M | B/R | BCNCS | BCNCS | Absent | Hypoplasia |
| 7-12/M | L/L | / | BCNCS | - | - |
| 7-12/M | L/L | / | BCNCS | - | - |
| 7-12/F | L/L | / | BCNCS | - | - |
| 13-18/F | B/B | - | - | Hypoplasia | Hypoplasia |
| 13-18/M | B/R | BCNCS | BCNCS | - | - |
| 13-18/M | L/L | / | BCNCS and Vestibular<br>semicircular canal<br>deformity | - | - |
| 13-18/F | L/L | / | BCNCS | / | Absent |
| 13-18/F | B/B | BCNCS | BCNCS | Hypoplasia | Hypoplasia |
| 13-18/F | L/L | / | BCNCS | / | Absent |
| 19-24/F | B/B | - | - | Hypoplasia | Hypoplasia |
| 19-24/F | B/B | BCNCS | BCNCS | Hypoplasia | Hypoplasia |
| 25-30/F | R/R | BCNCS | / | - | - |
| 25-30/M | B/L | BCNCS | BCNCS | Absent | Absent |

PHL profound hearing loss, CN cochlear nerve, M male, F female, R right, L left, B both sides, BCNCS bony cochlear nerve canal stenosis, / Not evaluated, - no data, IP-I
Incomplete partition type I. Only ears with PHL were evaluated.

### DPOAEs and ASSR thresholds

The CND ears were further divided into the IV’ wave present and IV’ wave absent group. There were 47 DPOAE and 45 ASSR recordings in the IV’ wave present group, while in the IV’ wave absent group, there were 68 DPOAE and 58 ASSR recordings. The averaged DPOAE SPL in the IV’ wave present group was - 7.29 ± 5.515 dB, while averaged DPOAE SPL in the IV’ wave absent group was -7.57 ± 6.190 dB. Two independent samples t test showed no significant difference between the two groups (t = -0.250, p = 0.803). The DPOAEs of the two groups in different frequency were displayed in Figure 4.

**FIGURE 4.**
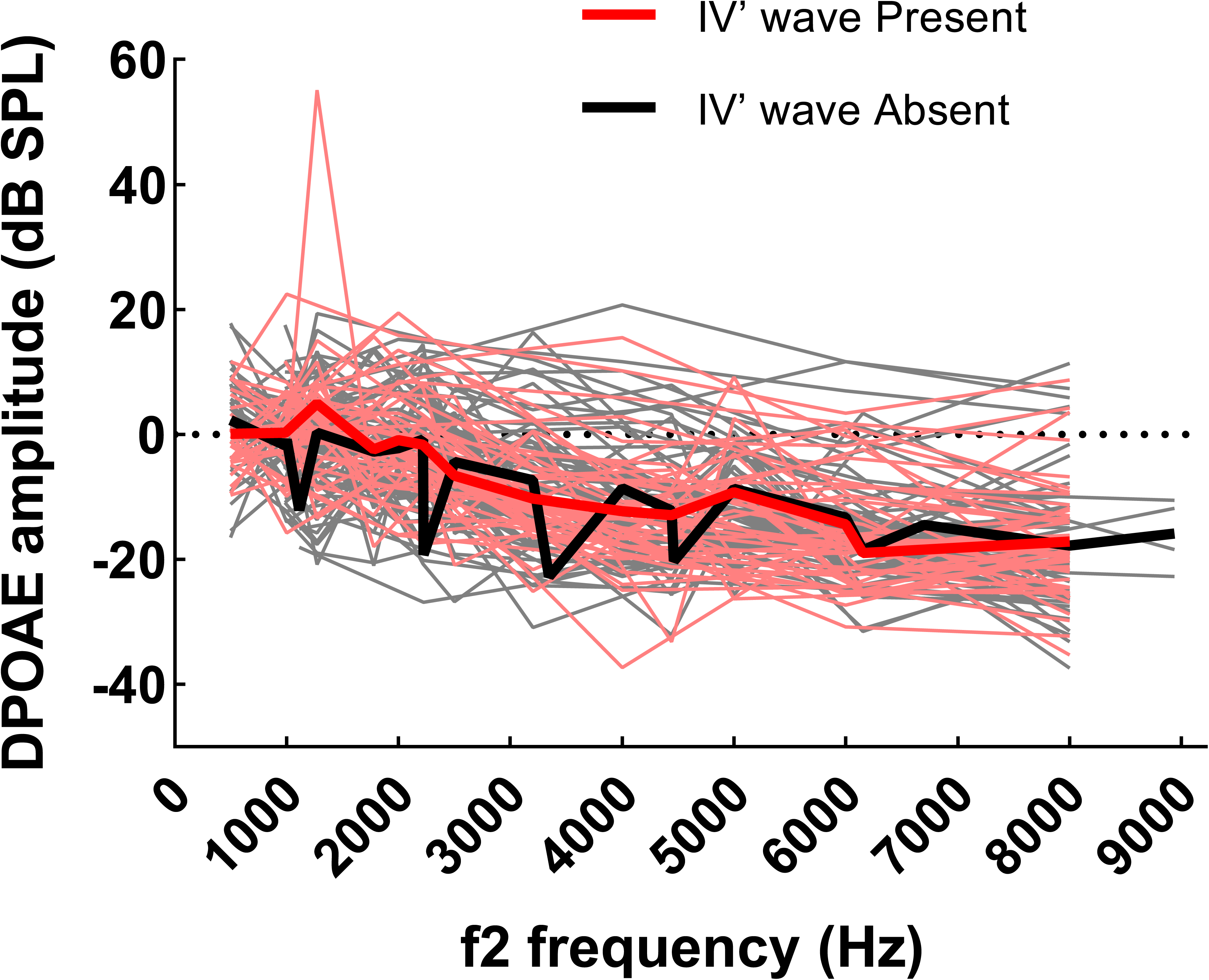
DPOAE absolute amplitude for the IV’ wave present and IV’ wave absent group. Individual (thin lines) and mean (thick lines) DPOAE absolute amplitude from 1 to 9 kHz for ears in IV’ wave present (red line) and IV’ wave absent group (gray line).

The results of extrapolated threshold of ASSR in the IV’ wave present and absent groups were displayed in Figure 5. The mean ± SD extrapolated threshold of frequency 500, 1000, 2000 and 4000 Hz is 80.7 ± 14.76, 82.6 ± 13.26, 84.7 ± 15.17 and 87.4 ± 19.99 dB HL in the IV’ wave present group. Those threshold in the IV’ wave absent group is 99.9 ± 21.47, 102.2 ± 15.11, 98.5 ± 17.82 and 103.5 ± 21.63 dB HL for frequency 500, 1000, 2000 and 4000 Hz respectively. Two independent samples t test showed that the difference was significant in each frequency between two groups (all p < 0.001).

**FIGURE 5.**
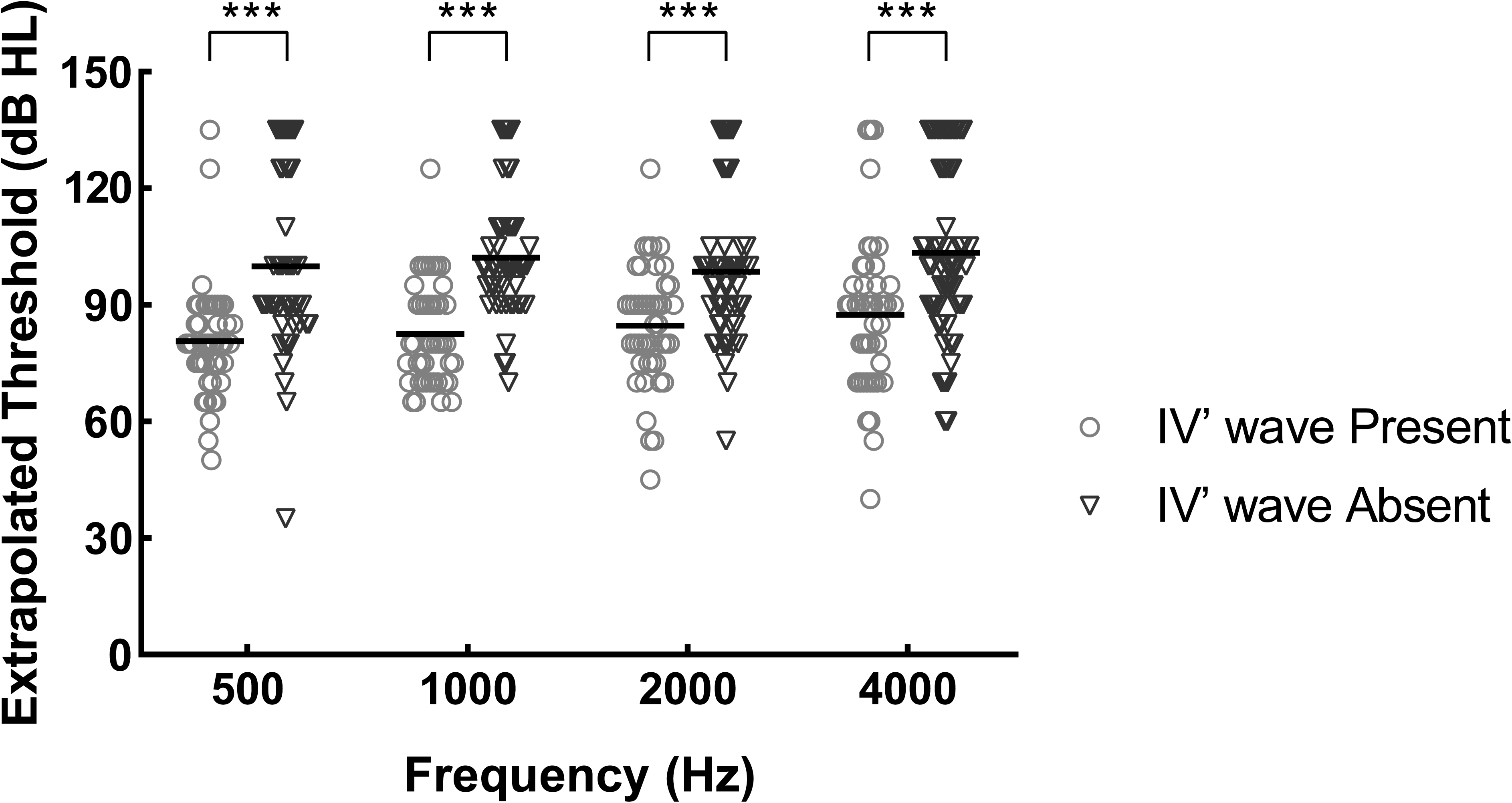
Individual and mean extrapolated threshold of ears in the IV’ wave present and IV’ wave absent group. *** indicates the difference was significant (p < 0.001)

### Follow-up of the IV’ wave

There are seven individuals presented with the IV’ wave at the first visit and had a follow-up visit later. The testing age and the IV’ wave display side of all seven individuals are listed in Table 2. The ABR recordings of the seven patients are displayed in Supplementary II. The follow-up period ranged from 3 to 11 months. The waveform morphology of the IV’ wave was slightly different depending on the different ABR recording systems (Case 01 and Case 07). However, in the same ABR recording system, the waveform morphology of the IV’ wave in the follow-up visit ABR recordings was similar to the previous waveforms. The IV’ wave was only barely observed in one individual 11 months later (Case 05).

**Table 2.**
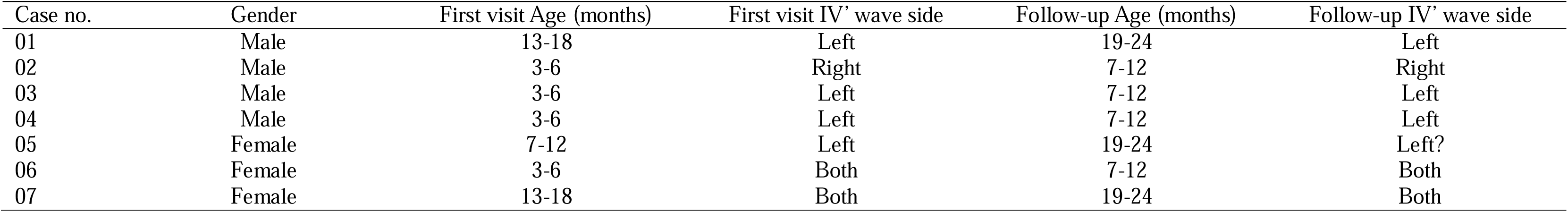
The testing age, gender and the IV’ wave display side of all seven individuals.

## DISCUSSION

In our study, we observed a 4.6 ms positive response, while the regular V wave was absent at the high intensity level in the ABR tests in some infants. The averaged latency of the IV’ wave at 95 dB is 4.60 ms. The positive predictive value of the IV’ wave for CND was 98.1%. The results of the extrapolated thresholds were significantly better in the IV’ wave present groups.

To the best of our knowledge, this is the first reported study of the special ABR waveform in infants with CND. ABRs are of high clinical relevance for objective analysis of hearing function, especially for screening for auditory neuropathy, acoustic neuroma, and central hearing loss^17,18^, as well as assessing “hidden hearing loss”^19–21^. In our study, the IV’ wave showed an excellent positive predictive value to CND and 39.5% of CND ears presented the IV’ wave. Additionally, the IV’ wave present group displayed better ASSR thresholds than those of the IV’ wave absent group. The results imply that the CN status is better in the IV’ wave present group and the IV’ wave is expected to be a new indicator in CND infants.

The outcomes of CI in patients with CND have been reported to be extremely variable^13,22–24^. Several authors have also stated that patients with CND have no chance to benefit from traditional CI and suggested direct stimulation of the cochlear nuclei by means of auditory brainstem implantation to improve auditory-verbal skills in such patients^25–27^. For the same reason, once the infants are diagnosed with unilateral CND, if the hearing of the other ear is within normal, CI is just an alternative. Many researchers are looking for objective predictive models for the outcome of CI in patients with CND^16,28–30^. However, all those indicators are based on imaging examinations. Freeman and Sennaroglu^31^ noted that radiological examination had a rate of false positives, and the absence of a vestibulocochlear nerve does not mean that the nerve is not present. Therefore, those radiological distractions could also have a negative impact on the predictive model. The IV’ wave specifically presented in the CND ears has the potential to be a new indicator for the CND population. The predictive accuracy of the models that applied both the IV’ wave and the radiological indicator could be improved effectively. In order to evaluate the predictive effect of the IV’ wave in CND patients, a series of studies are also needed in the future, including differences of the inner ear imaging and the outcome of the CI in the IV’ wave present CND patients.

Previous studies have shown that the ABR has a reasonably short maturational time course, the various peaks undergo changes in amplitude and latency at different rates in the first few years of life, and it does not become adult-like until around three years of age^32,33^. In our study, all ABR recordings were obtained in infants younger than three years old. The morphology profile of the IV’ wave was summed up. Most of the ABR recordings in the seven follow-up individuals presented the IV’ wave still. Whether the IV’ wave would be observed constantly and whether the latency and morphology would change in those infants until adulthood remains unknown. More studies of the IV’ wave in CND children and adults are needed in the future.

The origin of the IV’ wave remains unclear. The classic ABR is evoked by a 100-μs click of moderately high intensity level within 10ms after stimulus onset. The IV waves are invisible in normal click-ABR recordings sometimes. In contrast to the adult-like ABR, the infant and toddler ABR has slightly greater low-frequency energy. Whether the IV’ wave displayed in the infants in our study is the wave IV remains unclear. Wave IV is concluded to arises from midline brainstem structures, perhaps acoustic stria, trapezoid bodies, and the superior olivary complex^34^. Study has indicated a better identification of wave IV on the contralateral side with the bandpass filter setting at 300 to 3000 Hz^35^. However, in our study, the ABR was recorded ipsilaterally, the bandpass filter setting of the ABR test was 100 to 3000 Hz. The ABR wave of the other side that represent IV’ wave also varied. As a result, the obversion of the IV’ wave in our study is probably unconnected to the frequency of the bandpass filter. The multi-channel EEG research might shed some light on the origins of the IV’ wave in the future.

## CONCLUSION

Our study demonstrated a special ABR waveform with a positive response approximately 4.60 ms and an absent of regular V wave at the high intensity level in infants. The IV’ wave showed an excellent positive predictive value for CND. The ASSR results of the IV’ wave present group is significantly better than those of the IV’ wave absent group. The IV’ wave specifically presented in infants with CND is expected to be a new indicator for the CND population.

## Data Availability

All data produced in the present work are contained in the manuscript

## List of SDC

Supplemental Digital Content 1.docx

Supplemental Digital Content 2.pdf

## REFERENCES

1. Rance G. Auditory neuropathy/dys-synchrony and its perceptual consequences. Trends Amplif. 2005;9(1):1–43. doi:10.1177/108471380500900102

2. Amatuzzi MG, Northrop C, Liberman MC, et al. Selective inner hair cell loss in premature infants and cochlea pathological patterns from neonatal intensive care unit autopsies. Arch Otolaryngol Head Neck Surg. Jun 2001;127(6):629–36. doi:10.1001/archotol.127.6.629

3. Starr A, McPherson D, Patterson J, et al. Absence of both auditory evoked potentials and auditory percepts dependent on timing cues. Brain. Jun 1991;114 (Pt 3):1157–80. doi:10.1093/brain/114.3.1157

4. Starr A, Michalewski HJ, Zeng FG, et al. Pathology and physiology of auditory neuropathy with a novel mutation in the MPZ gene (Tyr145->Ser). Brain. Jul 2003;126(Pt 7):1604–19. doi:10.1093/brain/awg156

5. Shelton C, Luxford WM, Tonokawa LL, Lo WW, House WF. The narrow internal auditory canal in children: a contraindication to cochlear implants. Otolaryngol Head Neck Surg. Mar 1989;100(3):227–31. doi:10.1177/019459988910000310

6. Blegvad B, Hvidegaard T. Hereditary dysfunction of the brain stem auditory pathways as the major cause of speech retardation. Scand Audiol. 1983;12(3):179–87. doi:10.3109/01050398309076244

7. Lin PH, Hsu CJ, Lin YH, et al. An integrative approach for pediatric auditory neuropathy spectrum disorders: revisiting etiologies and exploring the prognostic utility of auditory steady-state response. Sci Rep. Jun 17 2020;10(1):9816. doi:10.1038/s41598-020-66877-y

8. Adunka OF, Jewells V, Buchman CA. Value of computed tomography in the evaluation of children with cochlear nerve deficiency. Otol Neurotol. Aug 2007;28(5):597–604. doi:10.1097/01.mao.0000281804.36574.72

9. Miyasaka M, Nosaka S, Morimoto N, Taiji H, Masaki H. CT and MR imaging for pediatric cochlear implantation: emphasis on the relationship between the cochlear nerve canal and the cochlear nerve. Pediatr Radiol. Sep 2010;40(9):1509–16. doi:10.1007/s00247-010-1609-7

10. Pagarkar W, Gunny R, Saunders DE, Yung W, Rajput K. The bony cochlear nerve canal in children with absent or hypoplastic cochlear nerves. Int J Pediatr Otorhinolaryngol. Jun 2011;75(6):764–73. doi:10.1016/j.ijporl.2011.02.017

11. Ren C, Lin Y, Xu Z, Fan X, Zhang X, Zha D. Audiological characteristics and cochlear implant outcome in children with cochlear nerve deficiency. Front Neurol. 2022;13:1080381. doi:10.3389/fneur.2022.1080381

12. Yousef M, Mesallam TA, Garadat SN, et al. Audiologic Outcome of Cochlear Implantation in Children With Cochlear Nerve Deficiency. Otol Neurotol. Jan 2021;42(1):38–46. doi:10.1097/MAO.0000000000002849

13. Degirmenci Uzun E, Batuk MO, D’Alessandro HD, Sennaroglu G. Auditory perception in pediatric cochlear implant users with cochlear nerve hypoplasia. Int J Pediatr Otorhinolaryngol. Sep 2022;160:111248. doi:10.1016/j.ijporl.2022.111248

14. Kutz JW, Jr., Lee KH, Isaacson B, Booth TN, Sweeney MH, Roland PS. Cochlear implantation in children with cochlear nerve absence or deficiency. Otol Neurotol. Aug 2011;32(6):956–61. doi:10.1097/MAO.0b013e31821f473b

15. Kari E, Gillard DM, Chuang N, Go JL. Can Imaging Predict Hearing Outcomes in Children With Cochleovestibular Nerve Abnormalities? Laryngoscope. Jun 2022;132 Suppl 8:S1–S15. doi:10.1002/lary.30008

16. Lu S, Xie J, Wei X, et al. Machine Learning-Based Prediction of the Outcomes of Cochlear Implantation in Patients With Cochlear Nerve Deficiency and Normal Cochlea: A 2-Year Follow-Up of 70 Children. Front Neurosci. 2022;16:895560. doi:10.3389/fnins.2022.895560

17. Roeser RJ, Valente M, Hosforddunn H. Audiology Diagnosis. 2007;

18. Lewis JD, Kopun J, Neely ST, Schmid KK, Gorga MPJJotASoA. Tone-burst auditory brainstem response wave V latencies in normal-hearing and hearing-impaired ears. 2015;138(5):3210.

19. Kujawa SG, Liberman MCJJoNtOJotSfN. Adding Insult to Injury: Cochlear Nerve Degeneration after "Temporary" Noise-Induced Hearing Loss. 2009;29(45):14077.

20. Mehraei G, Hickox AE, Bharadwaj HM, et al. Auditory brainstem response latency in noise as a marker of cochlear synaptopathy. 2016;36(13)

21. Ridley CL, Kopun JG, Neely ST, Gorga MP, Rasetshwane DM. Using Thresholds in Noise to Identify Hidden Hearing Loss in Humans. Ear Hear. Sep/Oct 2018;39(5):829–844. doi:10.1097/AUD.0000000000000543

22. Zhang Z, Li Y, Hu L, Wang Z, Huang Q, Wu H. Cochlear implantation in children with cochlear nerve deficiency: a report of nine cases. Int J Pediatr Otorhinolaryngol. Aug 2012;76(8):1188–95. doi:10.1016/j.ijporl.2012.05.003

23. Young NM, Kim FM, Ryan ME, Tournis E, Yaras S. Pediatric cochlear implantation of children with eighth nerve deficiency. Int J Pediatr Otorhinolaryngol. Oct 2012;76(10):1442–8. doi:10.1016/j.ijporl.2012.06.019

24. Wu CM, Lee LA, Chen CK, Chan KC, Tsou YT, Ng SH. Impact of cochlear nerve deficiency determined using 3-dimensional magnetic resonance imaging on hearing outcome in children with cochlear implants. Otol Neurotol. Jan 2015;36(1):14–21. doi:10.1097/MAO.0000000000000568

25. Colletti V, Fiorino F, Sacchetto L, Miorelli V, Carner M. Hearing habilitation with auditory brainstem implantation in two children with cochlear nerve aplasia. Int J Pediatr Otorhinolaryngol. Aug 20 2001;60(2):99–111. doi:10.1016/s0165-5876(01)00465-7

26. Sennaroglu L, Ziyal I, Atas A, et al. Preliminary results of auditory brainstem implantation in prelingually deaf children with inner ear malformations including severe stenosis of the cochlear aperture and aplasia of the cochlear nerve. Otol Neurotol. Sep 2009;30(6):708–15. doi:10.1097/MAO.0b013e3181b07d41

27. Buchman CA, Teagle HF, Roush PA, et al. Cochlear implantation in children with labyrinthine anomalies and cochlear nerve deficiency: implications for auditory brainstem implantation. Laryngoscope. Sep 2011;121(9):1979–88. doi:10.1002/lary.22032

28. Chung J, Jang JH, Chang SO, et al. Does the Width of the Bony Cochlear Nerve Canal Predict the Outcomes of Cochlear Implantation? Biomed Res Int. 2018;2018:5675848. doi:10.1155/2018/5675848

29. Han JJ, Suh MW, Park MK, Koo JW, Lee JH, Oh SH. A Predictive Model for Cochlear Implant Outcome in Children with Cochlear Nerve Deficiency. Sci Rep. Feb 4 2019;9(1):1154. doi:10.1038/s41598-018-37014-7

30. Birman CS, Powell HR, Gibson WP, Elliott EJ. Cochlear Implant Outcomes in Cochlea Nerve Aplasia and Hypoplasia. Otol Neurotol. Jun 2016;37(5):438–45. doi:10.1097/MAO.0000000000000997

31. Freeman SR, Sennaroglu L. Management of Cochlear Nerve Hypoplasia and Aplasia. Adv Otorhinolaryngol. 2018;81:81–92. doi:10.1159/000485542

32. Hecox K, Burkard R. Developmental dependencies of the human brainstem auditory evoked response. Ann N Y Acad Sci. 1982;388:538–56. doi:10.1111/j.1749-6632.1982.tb50815.x

33. Fria TJ, Doyle WJ. Maturation of the auditory brain stem response (ABR): additional perspectives. Ear Hear. Nov-Dec 1984;5(6):361–5. doi:10.1097/00003446-198411000-00008

