## Supplementary material for "A Positive Response around 4.6 ms in Auditory Brainstem Response in Cochlear Nerve Deficiency Infants": Supplemntal 1

**ABR Recording and Stimulus Parameters**

The ABR tests were performed with the OtoAccess (Eclipse, Interacoustics, Denmark), Neuro-Audio.NET (Neurosoft, Neurosoft, Russia) and ICS Chartr EP (Chartr EP, Otometrics, Denmark) system. Alternative clicks were made through inserted earphones. Inserted earphones IP30 were used for the Eclipse and Neurosoft system. Otometrics inserted earphones were used for the Chartr EP system. The active electrode was placed on the center of the forehead, the ground electrode on the nasal root, and the reference electrodes on the left and right mastoid processes. The skin was prepared for electrode placement with a mild abrasive to obtain electrode impedances under 5 kohm and interelectrode impedances of less than 2 kohm. Each ear was stimulated separately, and each click lasted 100 ms with a repetition rate of 37.1 pulses/s. The bandpass filter was set at 100-3000 Hz. A total of 1,024 sweeps were recorded. The time window was 20 ms for the OtoAccess，15 or 20 ms for Neuro-Audio.NET system and 18 ms for the ICS Chartr EP system. The amplitude scale was 200 nv for all systems. For those individuals with single side deafness, there is a white noise masking on the other side in the test.

**ASSR Recording and Stimulus Parameters**

The ASSR tests were performed after the ABR tests with the same system. The insert earphone and the recording electrodes are the same with those in the ABR tests.

For the OtoAccess system, the Child (90Hz) Insert phone model was chosen. The stimuli rate model was chosen the child (90Hz). The stimuli consisted of NB CB-Chirp of four carrier frequencies (500, 1,000, 2,000 and 4,000 Hz) for the left and right ear respectively. The artificial level was set 160 μv. For those individuals with single side deafness, there is a white noise masking on the other side in the test.

For the Neuro-Audio.NET system, the child (90Hz) model was chosen. The stimuli of multiple ASSR consists Frequency-Specific Chirp of four carrier frequencies (500, 1,000, 2,000 and 4,000 Hz) with modulation at a rate of 90 Hz. The rejection level was set ± 18 μv. For those individuals with single side deafness, there is a white noise masking on the other side in the test.

For the ICS Chartr EP system, the Child Asleep Search model was chosen. The stimuli consisted of four carrier frequencies (500, 1,000, 2,000 and 4,000 Hz) with amplitude modulation at a rate of 90, 82, 98 and 94 Hz for the left ear stimuli and 88, 80, 96, and 92 for the right ear stimuli, respectively. The response confidence was set 95%. For those individuals with single side deafness, there is a masking of 80 dB HL on the other side in the test.

For all the systems, the response at each stimulation frequency were measured automatically; no subjective judgment by an interpreter was needed.

**DPOAE Recording and Stimulus Parameters**

Three commercially available DPOAE systems (OtoAccess, Neuro-Audio.NET and Capella) were used in the measurement and collection of DPOAE data. Before each use, the probe was calibrated by using the test option and a 2 cc acoustic calibration cavity. The equipment was set up with a f2/f1 ratio of 1.22 and stimulus frequency levels L1/L2 of 65/55 dB SPL. For the different DPOAE systems, the frequency of f2 is different. For the OtoAccess system, the f2 frequency is 500, 1000, 2000, 4000, 6000, and 8000 Hz. For the Neuro-Audio.NET system, the F2 frequency is 988, 1270, 1778, 2222, 2500, 3200, 4444, 5000, 6154 and 8000 Hz or 988, 1481, 2222, 4444, 5714 and 8000 Hz. For the Capella system, the F2 frequency is 1113, 2225, 3344, 4463, 6701 and 8939 Hz.
