## Supplementary material for "A Positive Response around 4.6 ms in Auditory Brainstem Response in Cochlear Nerve Deficiency Infants": Supplemntal 2

### Case 01

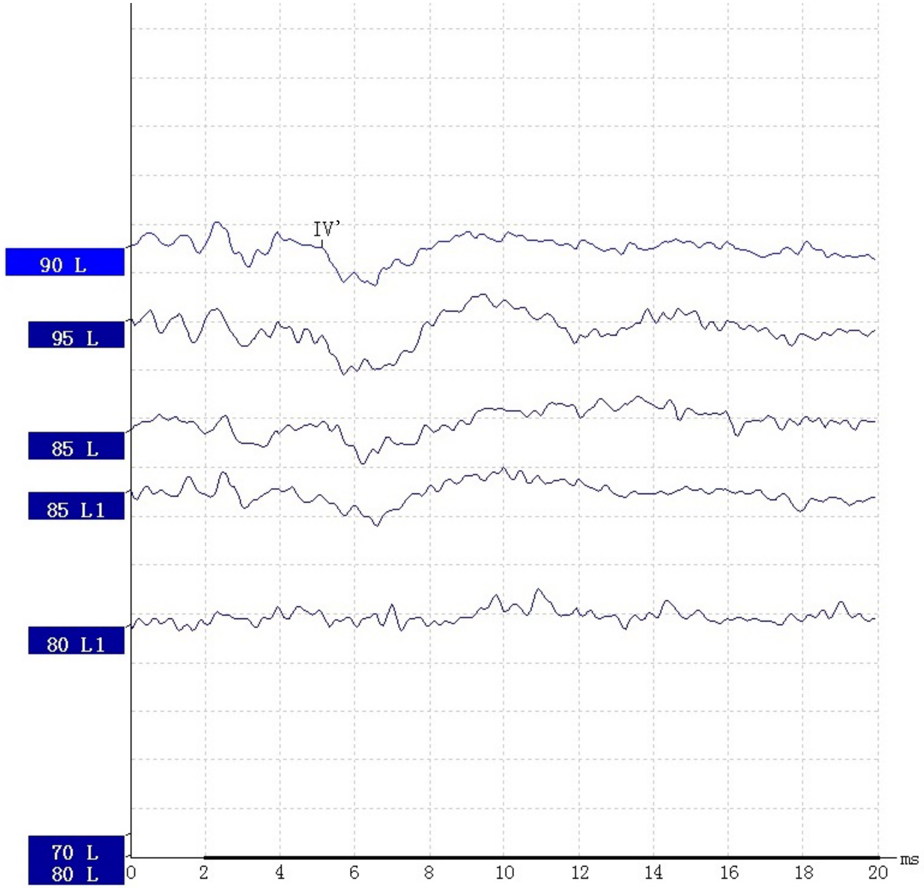


13-18 months

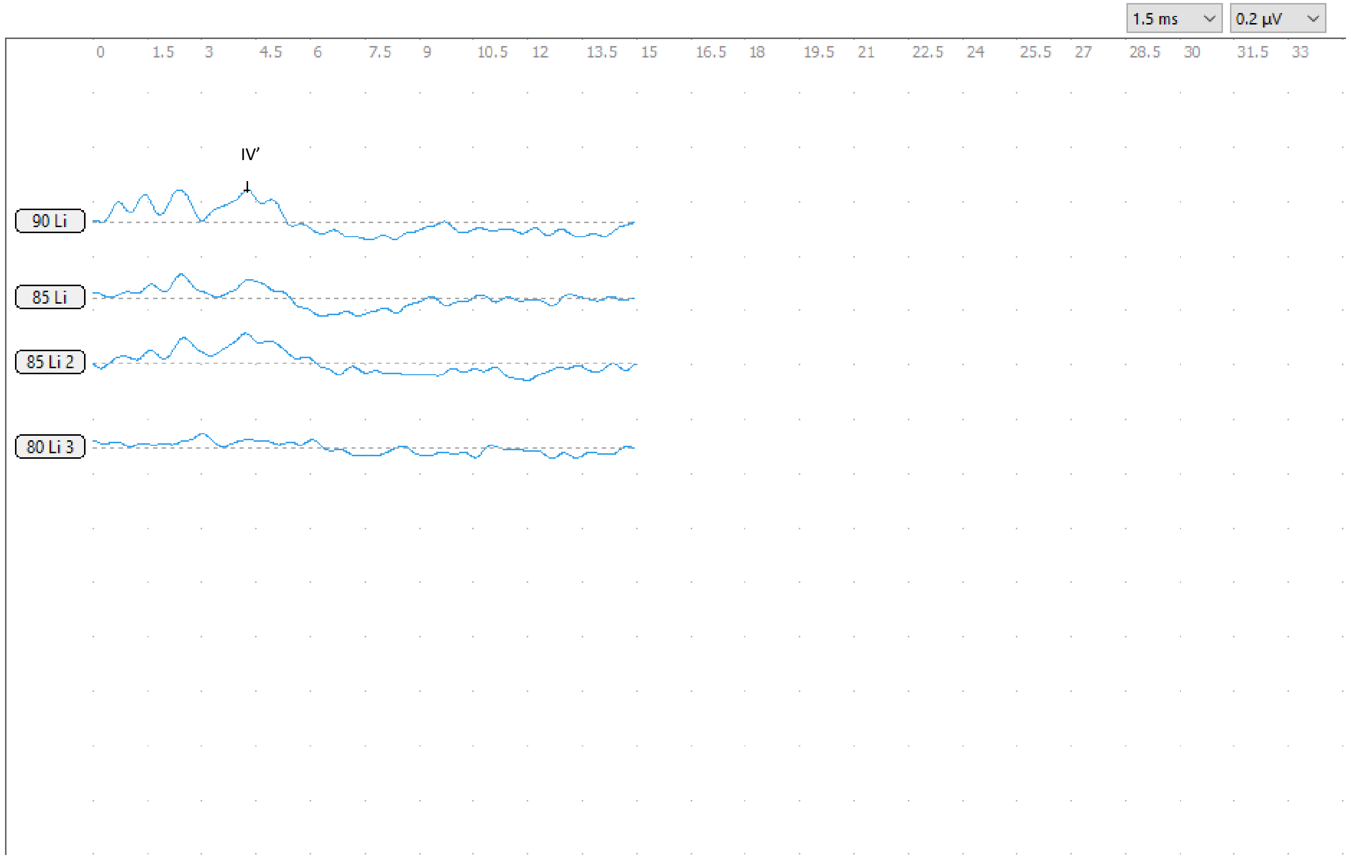


19-24 months

Case 02

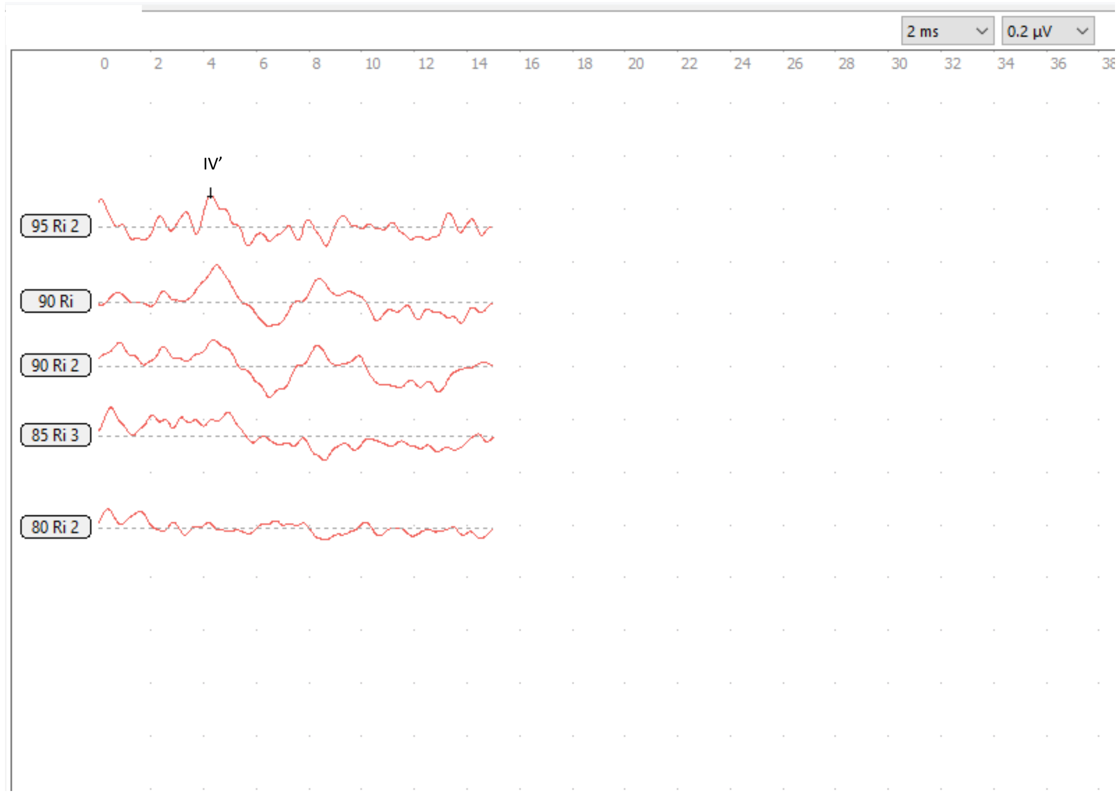


3-6 months

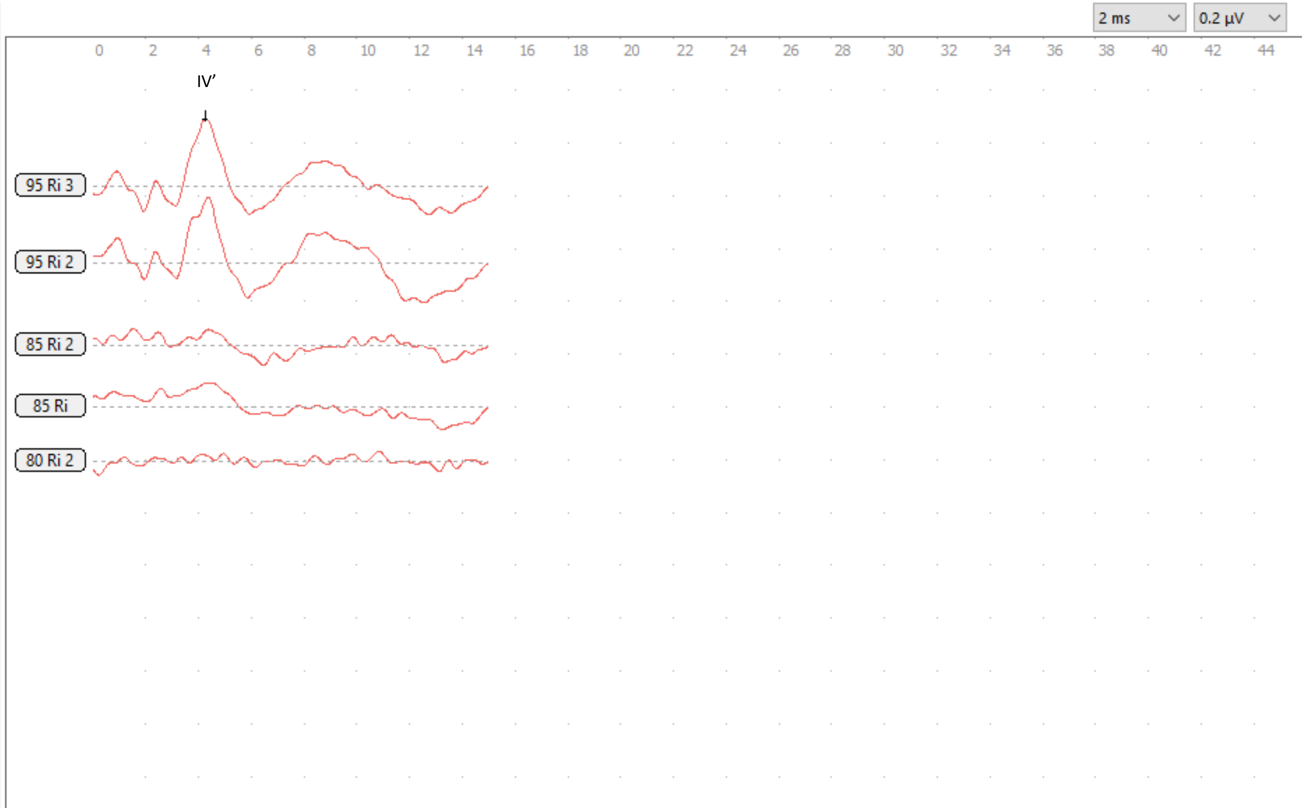


7-12 months

#### Case 03

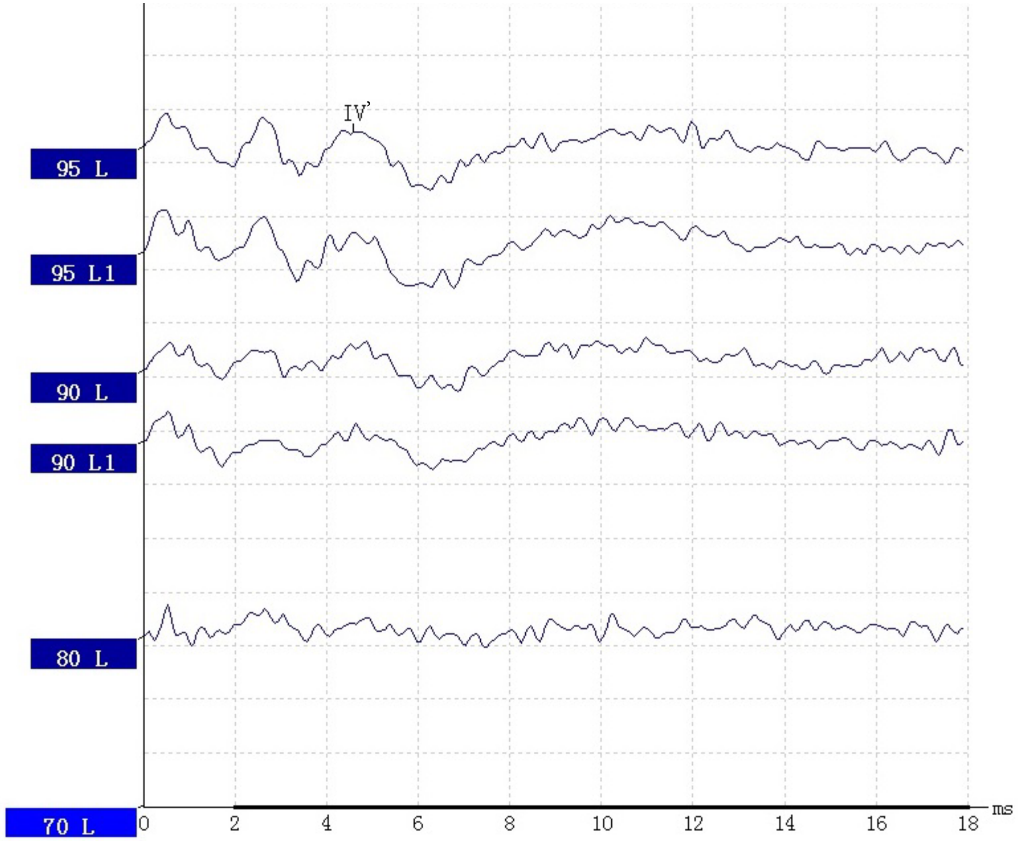


3-6 months

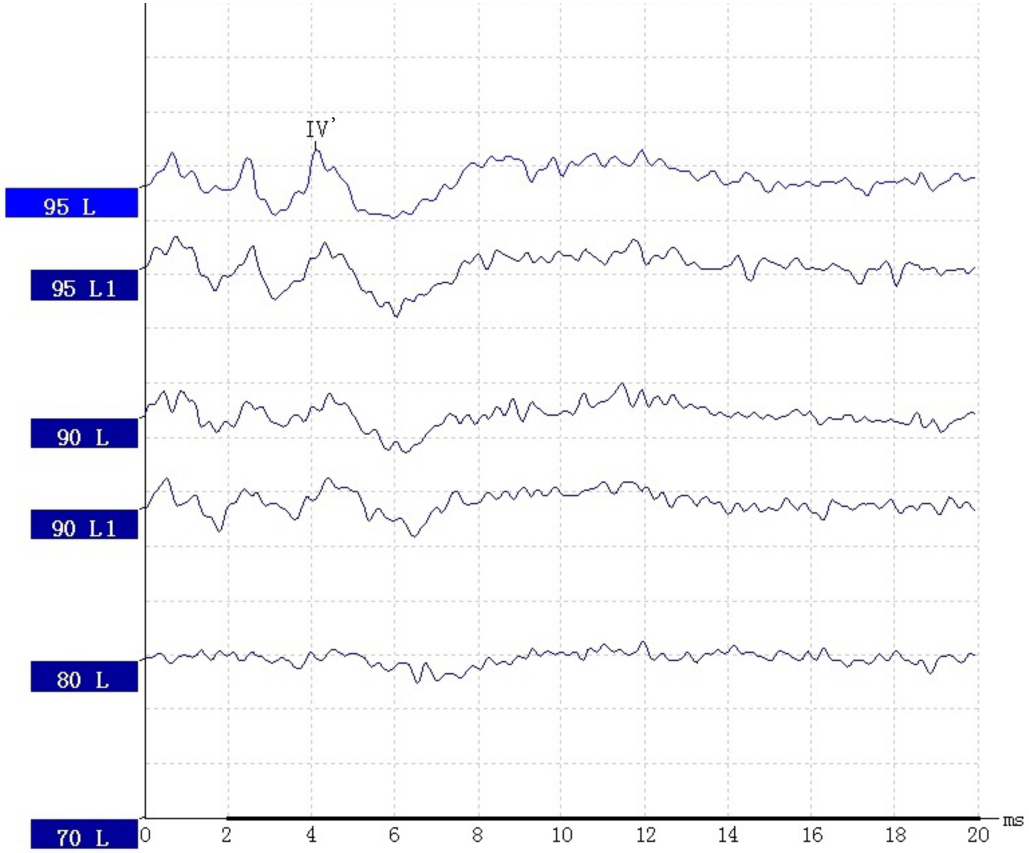


7-12 months

#### Case 04

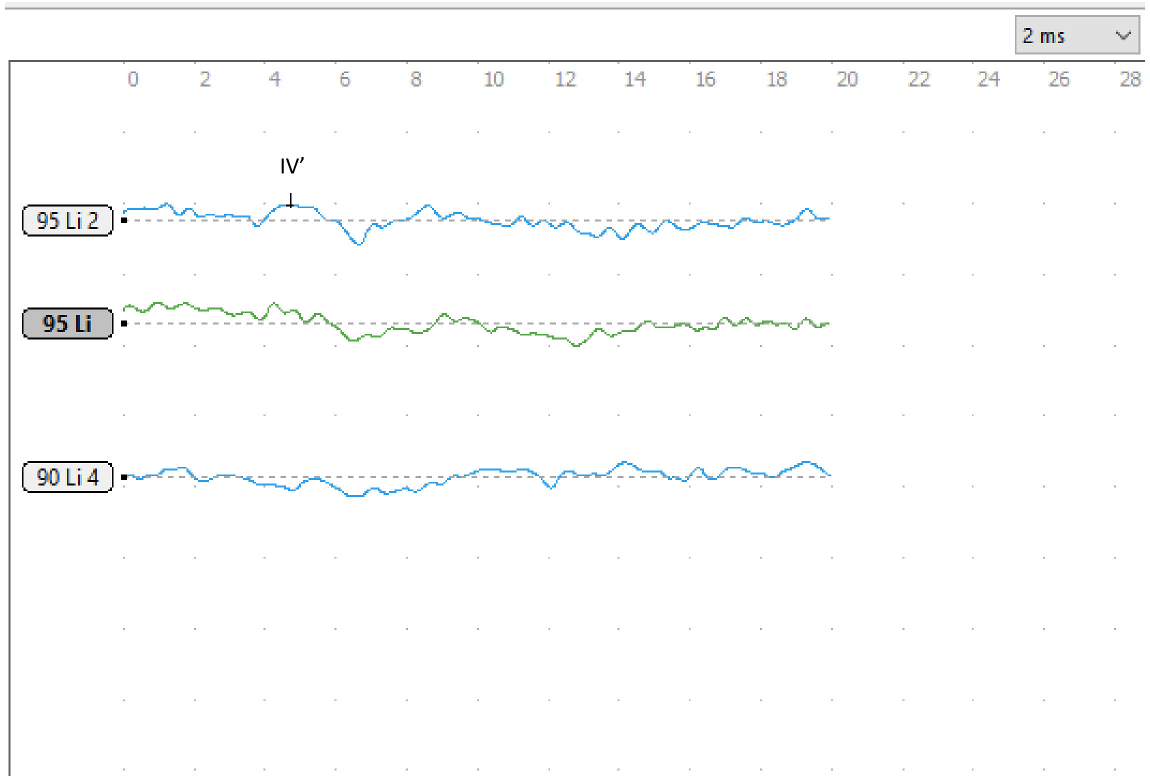


3-6 months

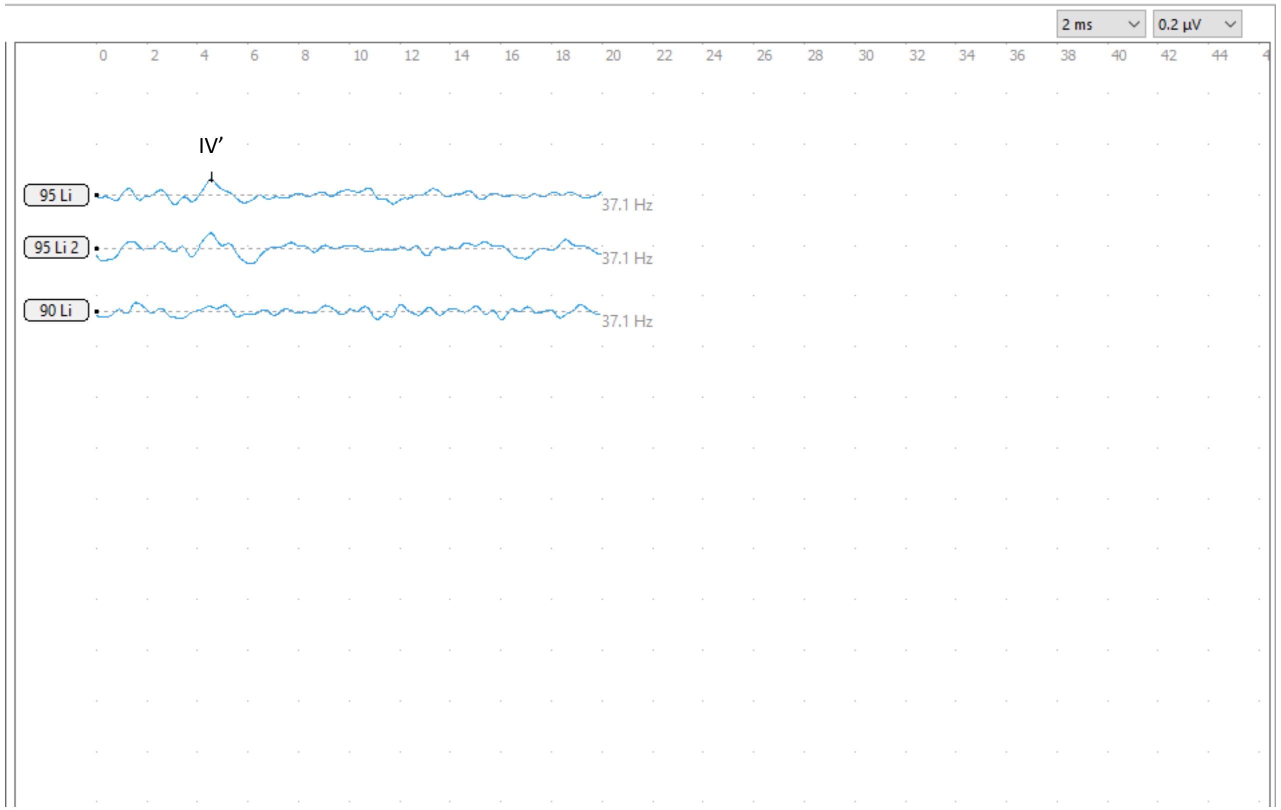


7-12 months

### Case 05

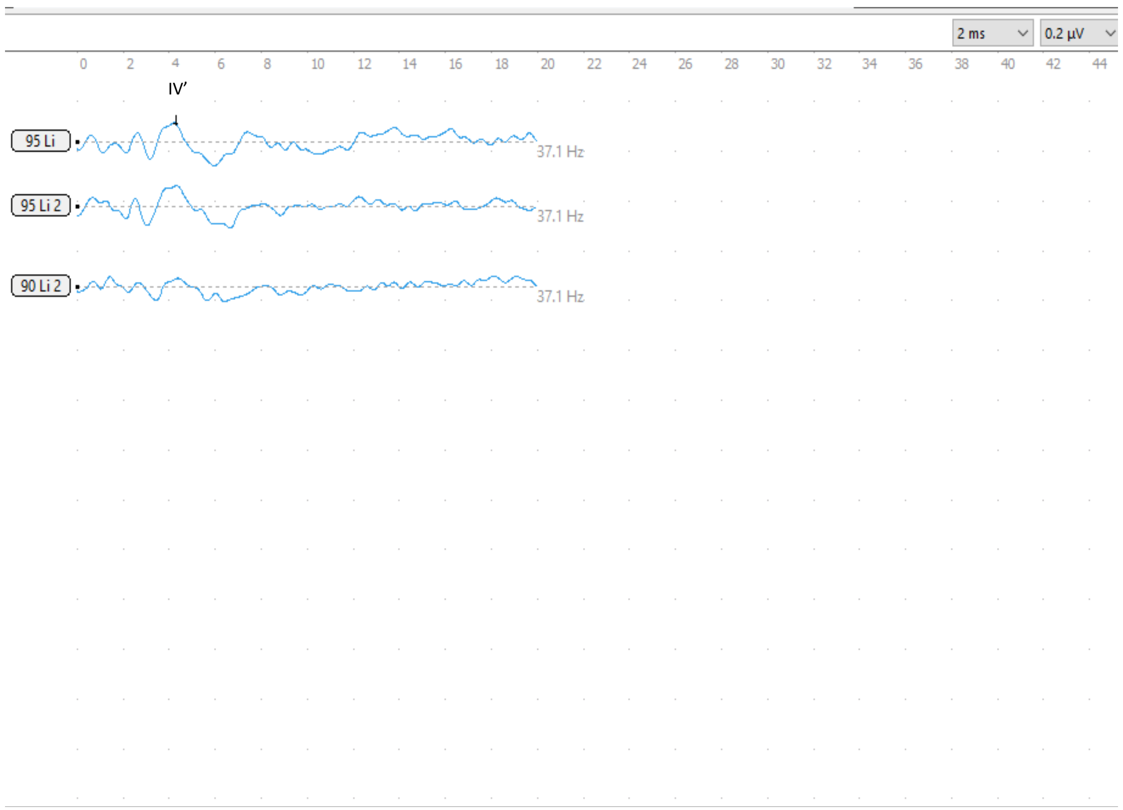


7-12 months

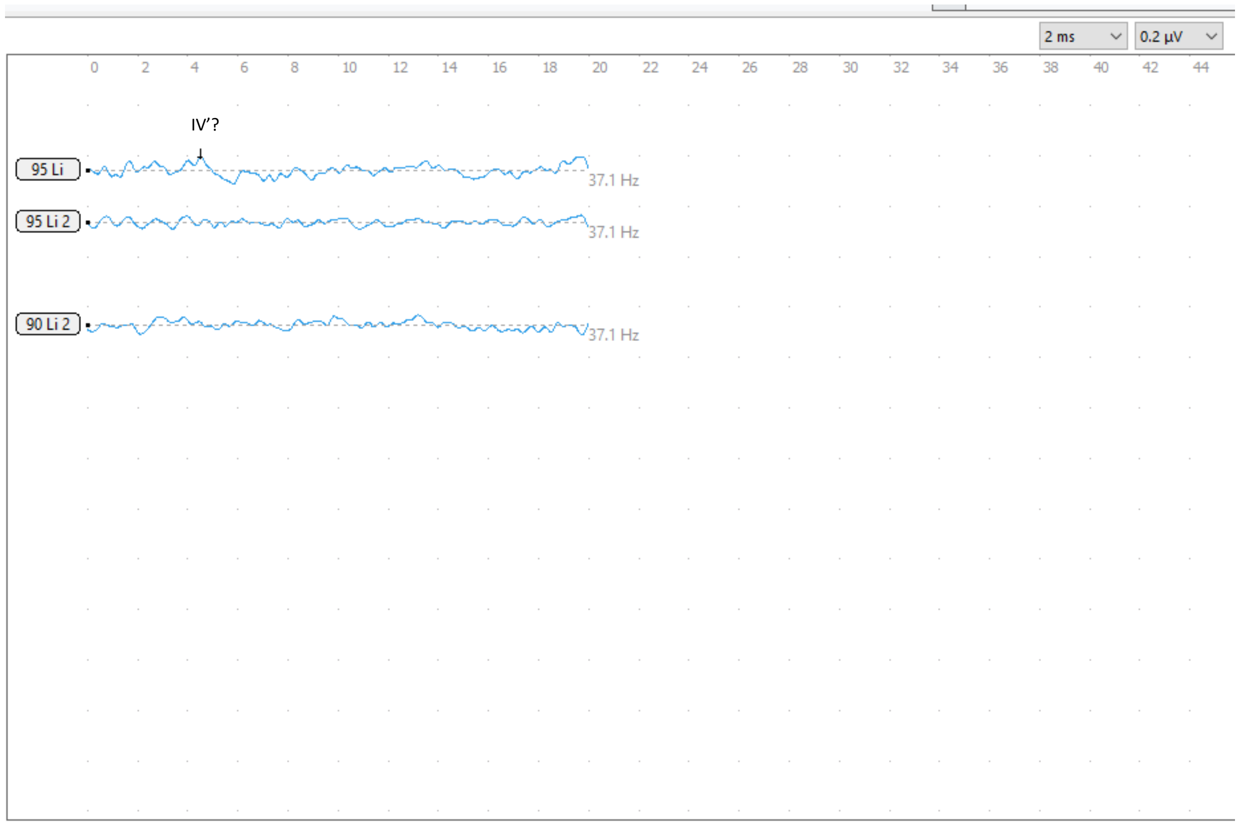


19-24 months

### Case 06

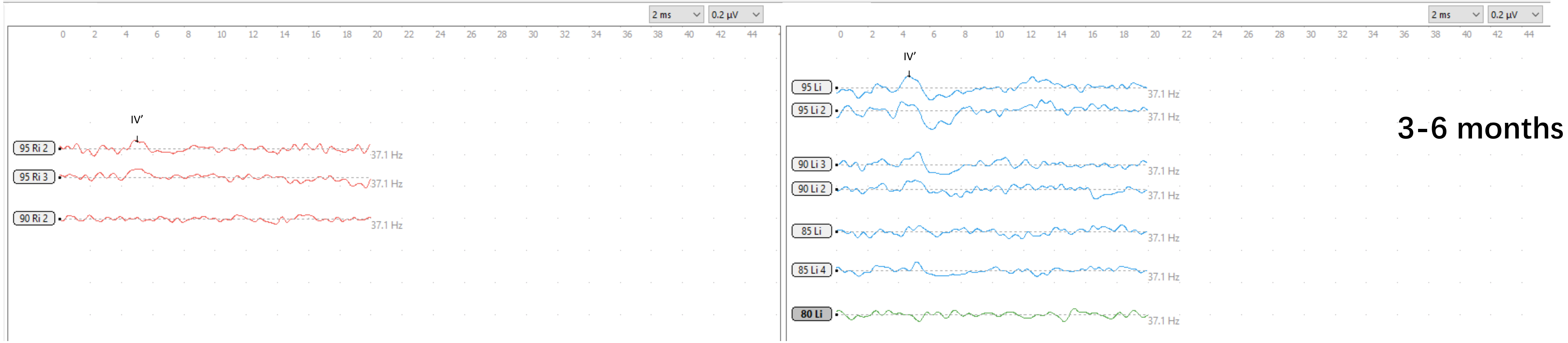

2 ms
0.2 µV
IV′
95 Ri 2
95 Ri 3
90 Ri 2
37.1 Hz
IV′
95 Li
95 Li 2
90 Li 3
90 Li 2
85 Li
85 Li 4
80 Li
3-6 months


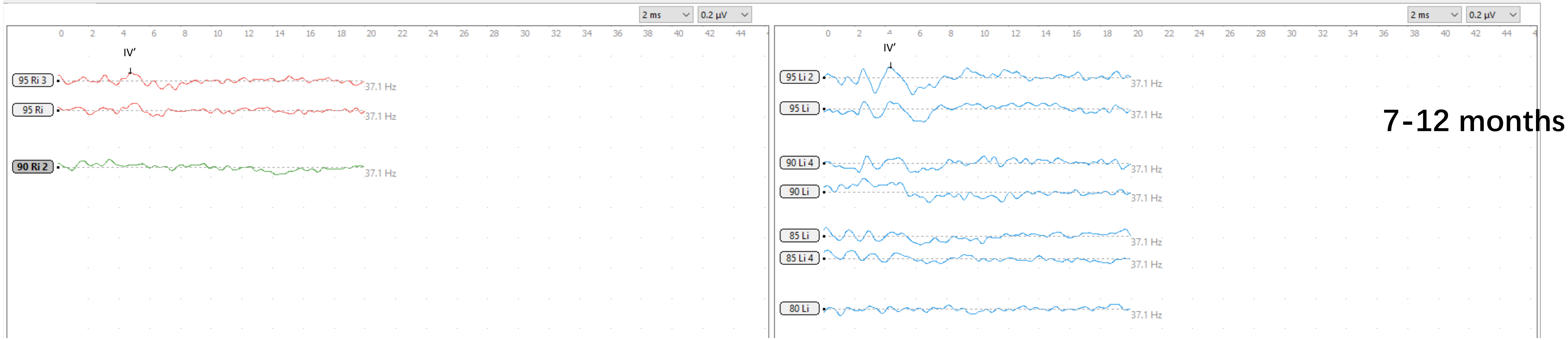

2 ms
0.2 µV
IV′
95 Ri 3
95 Ri
90 Ri 2
37.1 Hz
IV′
95 Li 2
95 Li
90 Li 4
90 Li
85 Li
85 Li 4
80 Li
7-12 months

### Case 07

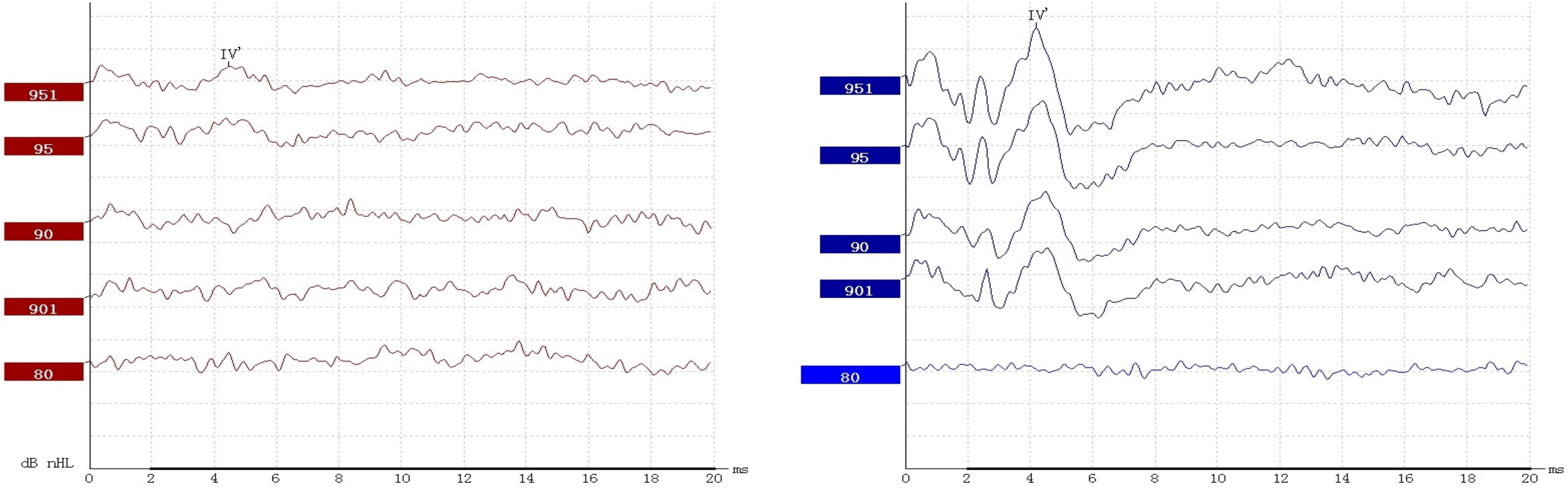


13-18 months

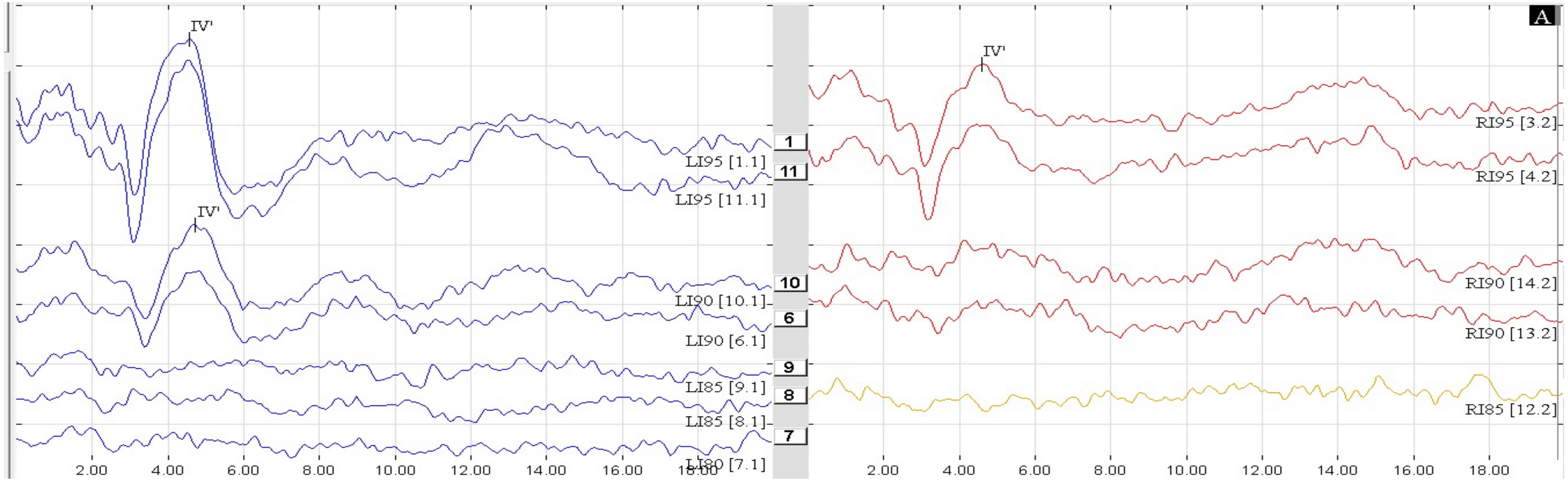


19-24 months
